# Measuring stroke outcomes using linked administrative data: Population-based estimates and validation of “home-time” as a surrogate measure of functional status

**DOI:** 10.1101/19005082

**Authors:** Melina Gattellari, Chris Goumas, Bin Jalaludin, John Worthington

## Abstract

**Background:** Administrative data offer cost-effective, whole-of-population stroke surveillance yet the lack of validated outcomes is a short-coming. The number of days spent living at home after stroke (“home-time”) is a patient-centred outcome that can be objectively ascertained from administrative data. Population-based validation against both severity and outcome measures and for all subtypes is lacking.

**Methods:** Stroke hospitalisations from a state-wide census in New South Wales, Australia, from July 1, 2005 to March 31, 2014 were linked to pre-hospital data, post-stroke admissions and deaths. We calculated correlations between 90-day home-time and Glasgow Coma Scale (GCS) scores, measured upon a patient’s initial contact with paramedics, and Functional Independence Measure (FIM) scores, measured upon entry to rehabilitation after the acute hospital stroke admission. Negative binomial regression models were used to identify predictors of home-time.

**Results:** Patients with stroke (N=74,501) spent a median of 53 days living at home after the event. Median home-time was 60 days after ischaemic stroke, 49 days after subarachnoid haemorrhage and 0 days after intracerebral haemorrhage. GCS and FIM scores significantly correlated with home-time (p-values<0.001). Female sex predicted less home-time in ischaemic stroke, while being married increased home time after ischaemic stroke and subarachnoid haemorrhage.

**Conclusions:** Home-time measured using administrative data is a robust, replicable and valid patient-centred outcome enabling inexpensive population-based surveillance.

## 1. Introduction

Stroke is a leading cause of death and disability with significant global disparities in incidence, mortality and morbidity [1]. Over the past two decades there have been considerable advances in primary and secondary stroke prevention, particularly in high-income nations [2-5] and, more recently, health services have been re-designed in several countries to facilitate access to multidisciplinary stroke unit care, thrombolysis and mechanical thrombectomy for ischaemic stroke and endovascular coiling for subarachnoid haemorrhage [6-9]. Monitoring stroke outcomes can assist in identifying areas of need, informing implementation strategies to bridge evidence-practice gaps. Administrative data offer a cost-effective alternative to labour-intensive gold-standard epidemiological studies, providing whole-of-population stroke surveillance [10]. However, the absence of validated outcomes after stroke is a significant shortcoming.

Home-time measures the number of days spent at home after stroke onset, taking into account patient survival and days spent in hospital, reflecting a patient’s capacity to return to their private residence after hospitalisation [11-15]. Home-time therefore can be unobtrusively and objectively measured using routinely collected data that are linked to health records following stroke and is proposed as a patient-centred surrogate measure of functional status. Unlike traditional stroke outcome measures, such as the modified Rankin Scale, the validity of home-time is not threatened by inter-rater variability, attrition of research participants or respondent characteristics (proxy versus patient) [16,17]. Home-time derived from administrative data may therefore be a cost-effective adjunct to traditional patient reported outcomes to minimise selection bias, improve the reliability of outcome assessment and allow continuous large-scale monitoring that otherwise would be impracticable. Deriving home-time from administrative data may also broaden patient representation in epidemiological and health services research. For example, patients whose preferred language is not the dominant community language or those with cognitive impairment may encounter barriers to effective engagement in research and quality assurance activities. Characterised in this way, home-time is inclusive, ascertained for all patients irrespective of their socio-demographic and health characteristics.

Few studies validating home-time are population-based or have validated home-time against both severity and outcome measures and for all stroke sub-types [11,14]. Further, there is limited information about prognostic indicators of home-time and validation has yet to be conducted where severity, home-time and functional status are measured wholly using routinely collected data. We analysed a jurisdiction-wide analysis reporting population-based estimates of home-time and validated home-time as an outcome measure for routine stroke surveillance.

## 2. Methods

Reporting according to the “**RE**porting of studies **C**onducted using **O**bservational **R**outinely-collected health **D**ata (RECORD) Statement” is reported in Supplementary Table 1.

### 2.1 Patient Selection

The study was part of the Home to Outcomes (H_2_0) project which aimed to report stroke epidemiology and outcomes in New South Wales (NSW), Australia’s most populous state (population ∼7.99 million in 2019). The project utilised linked routinely collected pre-hospital, hospital, rehabilitation and mortality data. As previously described [18,19] deterministic and probabilistic linkages were performed by the Centre for Health Record Linkage (CHeReL) using gold-standard privacy preserving protocols and ensuring that the linkage error rate was no greater than 5 per 1,000. Patient identifiers, such as names, addresses, dates of birth and dates of health service delivery (for example, dates of admission) facilitated linkage. Quality assurance of the linkage process undertaken by the CHeReL has been described (http://www.cherel.org.au/quality-assurance).

Patients over the age of 15 years were identified from the Admitted Patient Data Collection, a census of all hospital separations in NSW recording the principal diagnosis and up to 49 secondary diagnoses using the International Classification of Diseases, version 10, Australian Modification (ICD-10AM) [20]. Applying ICD-10AM codes recommended to assign stroke sub-types [21,22], we selected admissions with a principal diagnosis of subarachnoid haemorrhage (ICD-10AM I60), intracerebral haemorrhage (ICD-10AM I61, I62.9) and ischaemic stroke (ICS-10AM I63, I64) from January1, 2005 to March 31^st^, 2014 providing a minimum follow-up of 90-days to June 30, 2014, the most recently available data at the time linkage was performed. Strokes recorded in secondary diagnostic positions were included if the primary diagnosis indicated an acute stroke event, where iatrogenic causes were identified or where the stroke was flagged as occurring in hospital. Where primary diagnoses indicated I64 or I62.9 codes (unspecified stroke and unspecified haemorrhagic stroke, respectively), we interrogated records for more specific stroke codes to assign stroke-subtype. Sub-type was defined in this manner for 2,417 or 2.5% of cases; otherwise, we adhered to recommendations to code these cases as ischaemic stroke (that is, I64) [21] or intracerebral haemorrhage (that is, I62.9) [22].

As previously reported [18,19], patients where ineligible if resident outside NSW or if cases had concomitantly recorded major head trauma (ICD-10AM S04, S06, S07, S08, S17, S18, S01.7, S02.0, S02.1, S02.6, S02.7, S03.0, S09.0, S09.2, S09, S05.2, S05.7) or primary or metastatic cerebral neoplasms (C71,C70.0,C70.9, C79.3,D33.0, D33.1, D33.2, D33.3, D33.9). We excluded cases whose initial stroke diagnosis was revised upon transfer to another hospital by the end of the next day after presentation as presumed misclassified cases. Early discharges home, defined as completion of care allowing discharge within the first 48 hours, were excluded to increase the specificity of case ascertainment consistent with previous approaches [23-25] and remove patients with early symptom resolution suggesting transient ischaemic attack or stroke mimics and cases with unrevised provisional stroke diagnoses. Patients transferred for care outside the state and other transfers in the absence of contiguous records were also excluded as missing hospitalisation data precluded accurate calculation of home-time. The first eligible stroke admission (“index admission”) for each patient during the study period was selected for analysis.

### 2.2 Calculation of 90-day home-time

In keeping with other research [14], home-time was defined as the sum of full days not spent in hospital in the 90 days after the stroke admission. For patients dying or newly discharged to a nursing home within 90-days, zero days were counted from the date of death or discharge, respectively. Ninety-day “home-time” was calculated by linking stroke cases to subsequent hospital records and the state death register to June 30, 2014. Hospital records incorporated the acute stroke admission including inter-hospital transfers, any contiguously recorded in-patient rehabilitation following the acute period of stay and all subsequent acute and sub-acute readmissions for any reason, including rehabilitation. Hospital encounters also included same-day admissions, which included out-patient rehabilitation.

Prior to applying eligibility and exclusion criteria, we undertook data cleaning for cases with illogical death record data to facilitate valid calculation of 90-day home-time. Specifically, there were 70 death records with duplicate records indicating more than one probable linkage to mortality data. We selected the record with information consistent with data recorded in the other available linked data-sets (for example, consistent age and sex between death and admissions records). There were a further 38 records with dates of death preceding hospital discharge by more than one day. Of these, eleven were deemed clerical errors and accepted as valid, while the remaining were considered erroneously linked death records.

### 2.3 Measures validating home-time

#### 2.31 Proxy stroke severity measurement

As described previously [19], we developed a proxy measure of stroke severity incorporating the Glasgow Coma Scale (GCS) and mode of arrival (ambulance or private transport) by linking admissions to pre-hospital data obtained from Ambulance NSW, the sole provider of emergency services for the state. Total GCS scores were obtained from the initial pre-hospital vital signs survey recorded at first contact with paramedics. Pre-determined cut-offs categorised patients as fully alert with a maximum score of 15 or with mild, moderate or severe levels of lowered consciousness [26,27]. Self-presenting patients arriving to hospital by private transport were considered “ambulant”. Both arrival mode and GCS were near universally available, measured prospectively at first contact with health services and ascertained blinded to patient outcomes. The GCS and mode of arrival strongly predict stroke mortality [26-29], providing de-facto validity for the composite measure. Within the H_2_0 study cohort, we have published a validation of the proxy severity measure in ischaemic stroke (N>17,000 patients) demonstrating a strong association with 30-day mortality with cross-validation of findings (N>18,000 patients) [19].

#### 2.32 Functional Status

We obtained the 18-item Functional Independence Measure (FIM) [30] from the NSW Subacute and Non-Acute Data Collection, recorded upon patient’s first admission to inpatient rehabilitation within 90-days of the stroke admission. FIM scores were available from July 1, 2009 for the sub-set of publicly funded hospitals, which provided ∼70% of inpatient stroke rehabilitation in Australia during the study period [31]. The FIM comprises 19 items, with 13 measuring motor function and five measuring cognitive function. Each item was measured using a seven-point ordinal scale with higher scores indicating greater independence with everyday tasks, such as mobility, eating, bathing, memory and problem solving. Total scores ranged from 18 to 126, while motor and cognitive subscale scores ranged from 13 to 91 and 5 to 35, respectively.

### 2.4 Statistical analysis

As home-time had a skewed distribution, we report median values and the interquartile range (IQR) for all strokes and by stroke sub-type. Mean values (± Standard Deviation) were calculated to enable comparisons with other studies. We calculated Spearman correlations between home-time and GCS and FIM scores. These descriptive analyses were repeated excluding patients dying in hospital who, by definition, spent zero days at home. We also report mean and median values for “home-time” for each level of the proxy stroke severity measure incorporating the GCS categories and mode of arrival.

To identify predictors of home-time, multivariate negative binomial regression was carried out to take into account the skewed distribution of home-time and its measurement in counts of whole days. We used generalised estimation equations to account for inter-dependencies of data clustered on hospital of presentation. Modelling was carried out using PROC GENMOD (SAS 9.4, SAS institute Inc, Cary, USA) and an exchangeable working correlation. Adjusted means were derived from the model output. Potential predictors included age, sex, year of admission, marital status and our proxy stroke severity measure. In addition, we ascertained stroke history and Charlson comorbidities via linkage to prior hospitalisations up to July 1, 2001, providing 4.5 years of historical admissions data for all cases. To allow for departure from linearity, we included both linear and quadratic terms for age and modelled each Charlson comorbidity characterised by applying ICD-10 codes as reported by Quan, Sundararjan, Halfon et al [32]. Hemiplegia and cerebrovascular disease were removed as redundant [33] while the effect of HIV/AIDs was not assessed given its very low prevalence. We modelled a measure of socio-economic status, the Index of Relative Socio-economic Disadvantage (IRSD) [34] reflecting the degree of disadvantage within the geographical area of patient residence based on area-aggregated indicators such as income, educational status and occupation collected from a mandatory, national census carried out once every five years (>95% participation rate of eligible households) [35]. IRSD scores are categorised into quintile cut-offs with lower scores indicating greater relative disadvantage. Multivariate analyses also adjusted for the length of time patients spent in hospital in the twelve-months prior to the stroke admission to account for pre-existing comorbidity burden and health service utilisation proximal to the stroke event. Analyses were undertaken separately for each of the individual stroke sub-types given the well-established differences in patient characteristics, risk factors and prognosis. All analyses were carried out using SAS software (SAS 9.4, SAS institute Inc, Cary, USA).

### 2.5 Ethics

The NSW Population and Health Services Ethics Committee approved the study (HREC/14/CIPHS/17).

## 3. Results

### 3.1 Patient selection and characteristics

Of the 83,866 potentially eligible stroke admissions amongst NSW residents, there were 80,365 (95.8%) cases without known missing home-time data. Of these, 74,501 were index admissions including 57,949 patients with ischaemic stroke (77.8%), 12,192 with intracerebral haemorrhage (16.4%) and 4,360 with subarachnoid haemorrhage (5.9%) for which home-time was calculated (Figure 1). Almost half were female (N=37,097; 49.8%). One in four patients were older than 85 years of age (24.6%), while 7.0% were younger than 50 years (Supplementary Table 1).

**Figure 1:**
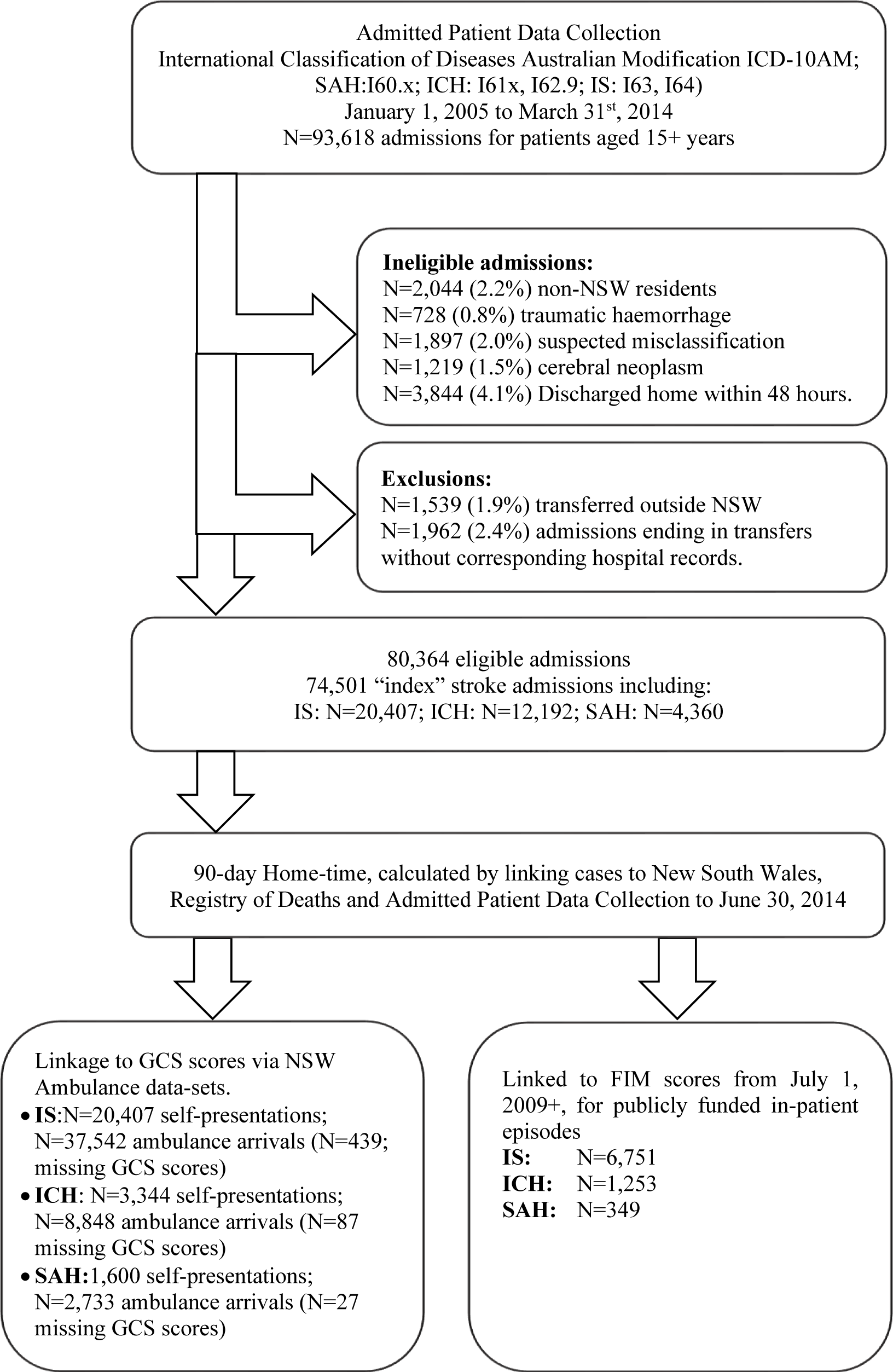
Linkage and patient selection flow chart.

### 3.2 Descriptive analysis of home-time estimates

Median home-time in the 90-days after stroke was 53 days (IQR=0 to 80 days). Median home-time was longer after an ischaemic stroke (Median=60-days; IQR=0 to 81) and shortest after an intracerebral haemorrhage (Median=0 days; IQR=0 to 63). Median home-time after a subarachnoid haemorrhage was 49 days (IQR=0 to 80 days). Almost 20% of patients (N=25,987) spent zero days at home in the 90-days after stroke. Just under one-third of ischaemic stroke patients (N=17,575; 30.3%) did not spend any full days at home compared with 55.2% (N=6,731) with intracerebral haemorrhage and 38.6% (N=1,681) with subarachnoid haemorrhage. When the study population was restricted to the sub-set of patients who survived their stroke admission, the estimated median home-time was similar for patients with ischaemic stroke and subarachnoid haemorrhage (Median=70 days and 69 days, respectively) and lower for those with an intracerebral haemorrhage (Median=55 days (IQR=5 to 78) (Table 1).

**Table 1.** Descriptive statistics for days at home “home-time”within 90 days of diagnosis by sex and age

| Sub-type | All cases |  |  | Excluding in-hospital deaths |  |  |
| --- | --- | --- | --- | --- | --- | --- |
|  | N | Mean (±SD) | Median (IQR) | N | Mean (±SD) | Median (IQR) |
| **Sub-type** |  |  |  |  |  |  |
| IS | 57,949 | 47±36 | 60 (0-81) | 48,153 | 56±32 | 70 (33-83) |
| ICH | 12,192 | 27±34 | 0 (0-63) | 7,147 | 46±33 | 55 (5-78) |
| SAH | 4,360 | 40±35 | 49 (0-74) | 3,082 | 56±28 | 69 (41-77) |
| *Total* | *74,501* | *43±36* | *53 (0-80)* | *58,382* | *55±32* | *69 (30-82)* |
| **Subtype by sex** |  |  |  |  |  |  |
| **IS** |  |  |  |  |  |  |
| Male | 29,653 | 51±35 | 66 (0-82) | 25,431 | 59±31 | 74 (41-83) |
| Female | 28,296 | 42±36 | 51 (0-80) | 22,722 | 53±33 | 66 (21-82) |
| **ICH** |  |  |  |  |  |  |
| Male | 6,059 | 29±35 | 0 (0-67) | 3,670 | 48±33 | 57 (12-80) |
| Female | 6,133 | 25±33 | 0 (0-59) | 3,477 | 43±33 | 52 (0-76) |
| **SAH** |  |  |  |  |  |  |
| Male | 1,692 | 42±35 | 55 (0-76) | 1,237 | 58±28 | 70 (43-78) |
| Female | 2,668 | 38±35 | 46 (0-73) | 1,845 | 55±29 | 68 (38-76) |
| **Subtype by age** |  |  |  |  |  |  |
| **IS** |  |  |  |  |  |  |
| <50 years | 2,925 | 66±28 | 80 (60-85) | 2,777 | 70±24 | 80 (67-85) |
| 50-64 years | 8,348 | 62±31 | 78 (46-84) | 7,775 | 66±27 | 79 (57-85) |
| 65-74 years | 11,668 | 55±33 | 72 (26-83) | 10,505 | 62±29 | 76 (47-84) |
| 75-84 years | 19,870 | 45±35 | 56 (0-80) | 16,471 | 54±32 | 67 (28-82) |
| 85+ years | 15,138 | 30±35 | 0 (0-67) | 10,625 | 43±34 | 52 (0-76) |
| **ICH** |  |  |  |  |  |  |
| <50 years | 967 | 44±36 | 54 (0-79) | 765 | 56±31 | 69 (35-82) |
| 50-64 years | 1,796 | 39±36 | 40 (0-78) | 1,282 | 54±31 | 66 (28-82) |
| 65-74 years | 2,350 | 32±35 | 8 (0-69) | 1,499 | 49±32 | 58 (17-81) |
| 75-84 years | 4,205 | 23±32 | 0 (0-53) | 2,277 | 43±33 | 49 (0-74) |
| 85+ years | 2,874 | 17±29 | 0 (0-28) | 1,324 | 36±33 | 38 (0-68) |
| **SAH** |  |  |  |  |  |  |
| <50 years | 1,313 | 53±32 | 69 (25-77) | 1,122 | 62±25 | 72 (56-78) |
| 50-64 years | 1,376 | 46±34 | 63 (0-75) | 1,096 | 58±27 | 69 (48-76) |
| 65-74 years | 674 | 34±34 | 25 (0-69) | 448 | 50±30 | 61 (25-76) |
| 75-84 years | 656 | 21±31 | 0 (0-48) | 311 | 44±31 | 51 (6-74) |
| 85+ years | 341 | 11±26 | 0 (0-0) | 105 | 37±35 | 39 (0-73) |

IS=Ischaemic Stroke
ICH=Intracerebral Haemorrhage
SAH=Subarachnoid Haemorrhage

**Table 2:**
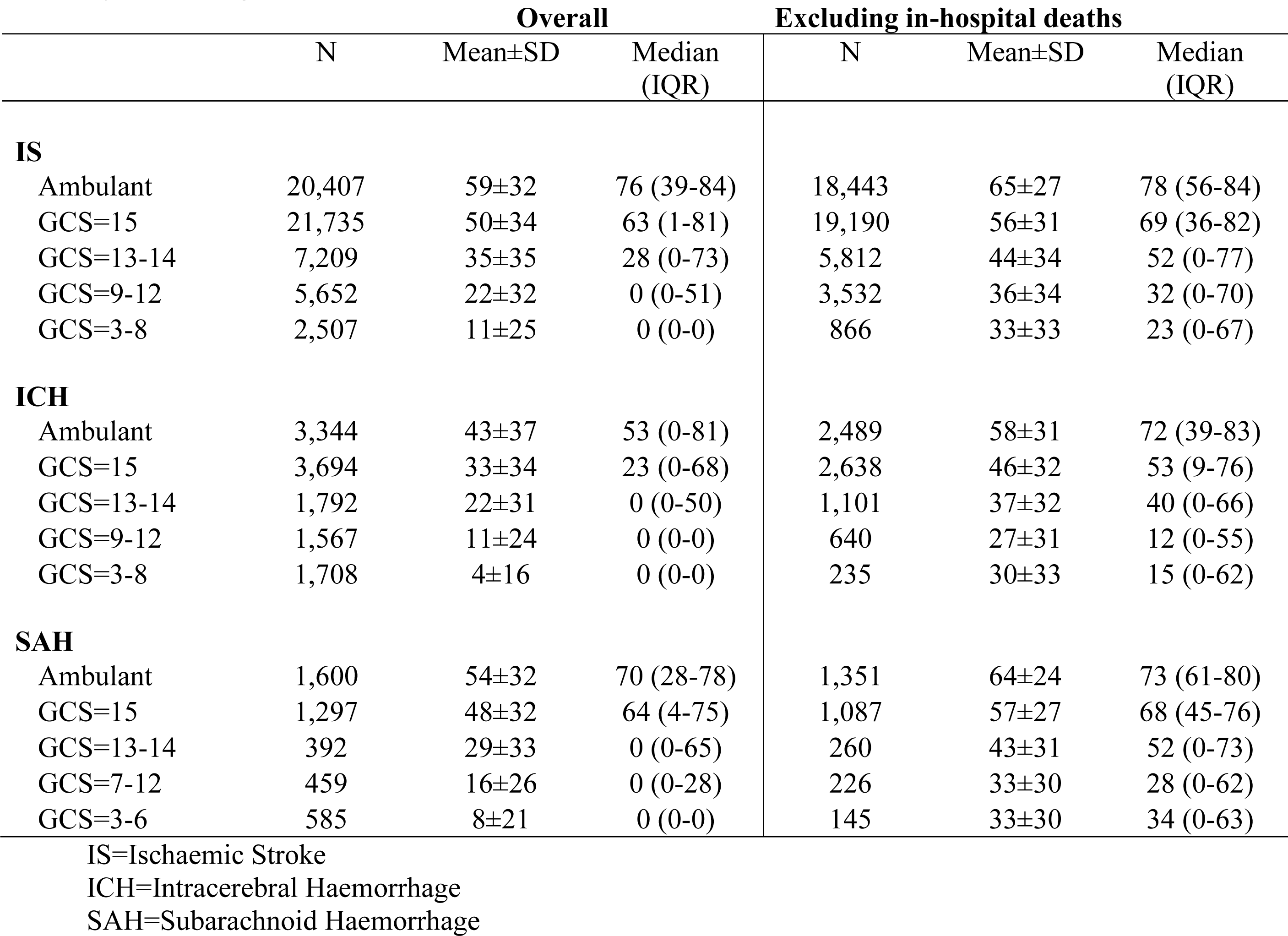
Descriptive statistics for days at home “home-time” within 90 days of diagnosis by sex and age

#### 3.2.1 Descriptive statistics for home-time according to age and sex

Median home-time was 15 days longer for males compared with females after an ischaemic stroke and nine days longer for men after a subarachnoid haemorrhage (Table 1). While the median home-time value for intracerebral haemorrhage was zero days for both sexes, the 75^th^ percentile value for home-time was 67 days for men and 59 days for women. When restricted to patients who survived their stroke admission at discharge, median values for males remained higher for all subtypes although differences were attenuated.

Increasing age was associated with a reduction in home-time for all sub-types. The decrease in median time across age was relatively gradual after ischaemic stroke ranging from 80 days in patients younger than 50 years of age to 56 days in those aged 75 to 84 years. Home-time reduced to zero days in patients older than 85 years. After intracerebral haemorrhage, median home-time decreased from 54 days in patients younger than 50 to 40 days in those aged 50-64 and then eight days in patients aged 65 to 74 years. After subarachnoid haemorrhage, median home-time decreased from 69 days in patients younger than 50 to 25 days in those aged 65-74 years. In both haemorrhagic stroke subtypes, median home-time was zero for patients older than 75 years.

### 3.3 Validity of home-time

#### 3.31 Association between home-time and proxy severity measure

Amongst self-presenting “ambulant” patients who did not require paramedic transport, the median home-time was 76 days (IQR=39-84) after ischaemic stroke, 53 days (IQR=0 to 81 days) after intracerebral haemorrhage and 70 days (IQR=28 to 78) after subarachnoid haemorrhage. GCS scores were available for 48,597 out of 49,150 (98.9%) patients transported via ambulance. After either an intracerebral or subarachnoid haemorrhage, median home-time was zero for patients with any decreased level of consciousness (GCS scores less than 15) (Table 1). Median home-time was zero after ischaemic stroke in patients with GCS scores less than 13. Amongst patients surviving their stroke admission, home-time decreased with decreasing levels of consciousness. However, in patients with haemorrhagic stroke, median home-time was higher for patients whose GCS scores indicated “very severe” brain injury (that is, unconscious patients), than for those with “severe” brain injury.

#### 3.32 Home-time and GCS and FIM scores

Higher GCS scores indicating higher levels of consciousness were moderately and significantly correlated with more home-time in ischaemic stroke and intracerebral haemorrhage (rho=0.35 and rho=0.39) (p<0.001) (Table 3). GCS scores more strongly correlated with home-time in subarachnoid haemorrhage (rho=0.51) (p<0.001). The correlation between GCS scores and home-time remained significant in the sub-group of patients who survived their stroke admission; however, the association was reduced (Table 3).

**Table 3:** Spearman correlation (rho) between GCS and FIM scores and “Home-Time”

| Sub-type | Overall |  | Excluding in-hospital deaths |  |
| --- | --- | --- | --- | --- |
|  | N | Rho | N | Rho |
| **GCS scores** |  |  |  |  |
| IS | 37,103 | 0.35 | 29,400 | 0.24 |
| ICH | 8,761 | 0.39 | 4,614 | 0.21 |
| SAH | 2,733 | 0.51 | 1,718 | 0.34 |
| **FIM scores** |  |  |  |  |
| Motor subscale |  |  |  |  |
| IS | 6,751 | 0.64 | 6,608 | 0.64 |
| ICH | 1,253 | 0.62 | 1,229 | 0.63 |
| SAH | 349 | 0.68 | 346 | 0.68 |
| Cognitive subscale |  |  |  |  |
| IS | 6,751 | 0.43 | 6,608 | 0.42 |
| ICH | 1,253 | 0.36 | 1,229 | 0.36 |
| SAH | 349 | 0.47 | 346 | 0.47 |
| Total FIM score |  |  |  |  |
| IS | 6,751 | 0.66 | 6,608 | 0.65 |
| ICH | 1,253 | 0.63 | 1,229 | 0.63 |
| SAH | 349 | 0.69 | 346 | 0.69 |

IS=Ischaemic Stroke
ICH=Intracerebral Haemorrhage
SAH=Subarachnoid Haemorrhage
All Spearman rho-values were statistically significant (p-values<0.001).

FIM scores were available from 2009 onwards for 8,353 patients undergoing publicly funded in-patient rehabilitation. FIM scores correlated with home-time for all sub-types (r_values_=0.63 to 0.69) (p-values<0.001). Motor scores were more strongly correlated with home-time than cognitive scores (r_values_=0.62 to 0.68 versus r_values_=0.36-0.47) (p-_values_<0.001) (Table 3) and were largely unchanged when restricted to patients surviving their stroke admission.

### 3.4 Predictors of home-time

Increasing stroke severity assessed using our proxy composite measure independently predicted fewer days spent at home after stroke for all subtypes in multivariate analyses (p-values<0.001) (Figure 2; Supplementary Tables 2-5).

**Figure 2:**
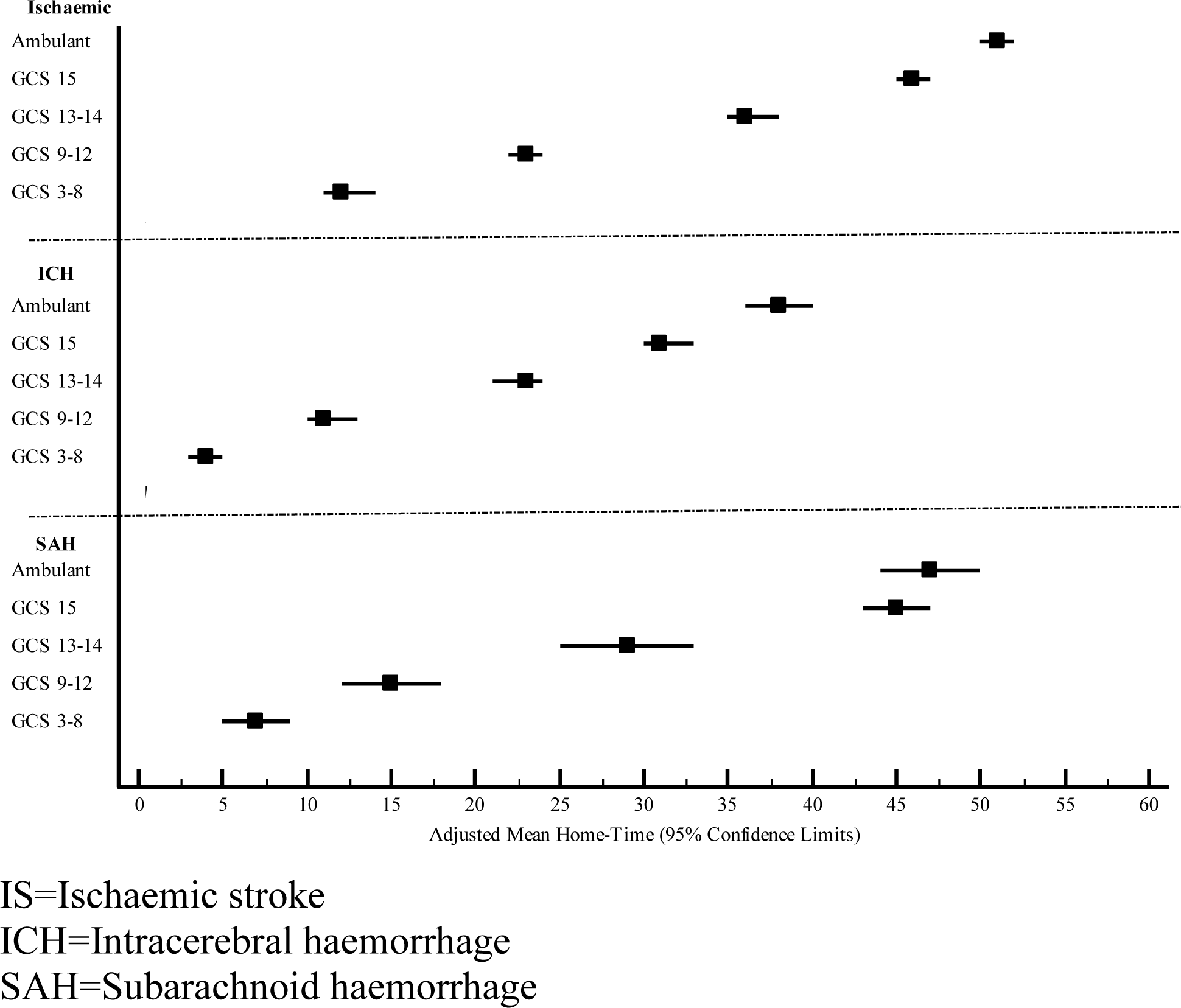
Adjusted Mean Home-time by stroke severity category.

Amongst ischaemic stroke patients, married patients spent significantly more days at home than single patients (Adjusted mean values=43 versus 39 days) (p<0.001) (Supplementary Table 3). The effect of socio-economic status was near significant (p=0.05). Patients residing in areas of least socio-economic disadvantage spent the greatest number of days at home (42 days) although differences in adjusted means between categories were small (one to two days). Home-time was reduced in the presence of all Charlson comorbidities tested, except connective tissue disease. Dementia (31 versus 41 days), metastatic malignancy (27 versus 41 days) and moderate or severe liver disease (30 versus 41 days) had the greatest impact on home time after ischaemic stroke. The adjusted effect of sex was significant (p=0.02). While adjusted mean home-time values for males and females were equivalent (41 days) the confidence intervals suggesting a higher range of estimated values for males (95% CI=41-42 versus 95% CI=40 to 41).

After intracerebral haemorrhage, patients spent significantly less time at home if they had a documented diagnosis of myocardial infarction (18 versus 21 days), congestive heart failure (17 versus 21 days), peripheral vascular disease (18 versus 21 days), renal disease (18 versus 21) and non-metastatic malignancy (17 versus 21 days). Patients with uncomplicated diabetes spent more time at home (23 versus 20 days) (Supplementary Table 4). Patients with subarachnoid haemorrhage spent significantly more time at home if they were married (32 versus 28 days) or if they had a previous stroke (40 versus 30 days) (p=0.02). The history of renal disease and moderate or severe liver disease were associated with less home-time after stroke (23 versus 31 days, p=0.03; 16 versus 31 days, p=0.03, respectively) (Supplementary Table 5). There was significant variation in home-time estimates according to year of admission for all sub-types, however, no clear linear association was evident.

## 4. Discussion

### 4.1 Main findings

Home-time was reduced with increasing age, more severe disease and after intracerebral and subarachnoid haemorrhage, all of which are known to adversely affect stroke outcome. Less functional independence and greater comorbidity burden also reduced home-time. These findings underscore the immediate adverse impact of stroke on people’s ability to live at home after the event and provide further evidence that home-time serves as a useful proxy for disability.

The large cohort afforded reliable insights into the impact of stroke in relatively young patients of which there were 5,205 cases aged younger than 50 years. Younger patients with intracerebral and subarachnoid haemorrhage spent less time at home compared with patients older than 75 after an ischaemic stroke. More than half of patients over the age 75 years after haemorrhagic stroke and half of those older than 85 years after an ischaemic stroke did not spend any time at home highlighting the considerable burden of stroke affecting elderly members of the community.

Females with ischaemic stroke spent fewer days at home as reported elsewhere [11,36]. While sex differences either narrowed or did not persist in multivariate analyses accounting for age, these findings demonstrate women’s vulnerability to poor outcomes due to their relatively advanced age, consistent with results from gold-standard incident stroke studies [37]. Patients residing in areas of greater socio-economic disadvantage and single patients with ischaemic stroke or subarachnoid haemorrhage had lower home-time estimates. Socio-demographic characteristics have also been associated with traditional stroke outcome measures [38]. These results either implicate patient characteristics in post-stroke morbidity or, alternatively, indicate that home-time measures constructs other than disability. For example, socio-demographic variables, may mediate their accessibility to health services post-stroke.

These findings inform the validity of the composite surrogate severity measure incorporating GCS and mode of arrival. The association between GCS and home-time was strongest for subarachnoid haemorrhage corroborating GCS as a valid component of standardised severity measures for this sub-type [27]. The correlation between GCS and home-time attenuated when in-patient deaths were removed from the cohort consistent with patterns of correlations reported when using gold-standard stroke severity assessment [14].

Previous results demonstrate that home-time correlates with the modified Rankin Scale or the Barthel Index [11-15,36,39]. Here, we demonstrate a strong correlation with another measure of functional status, the FIM, adding weight to the construct validity of home-time. Total FIM and motor subscale scores were strongly associated with home-time, while the correlation between cognitive subscale scores and days spent at home was more moderate. Home-time may therefore underestimate disability due to cognitive deficits, a shortcoming shared with traditional stroke outcome measures, such as the modified Rankin Scale score [40].

### 4.2 Comparison with previous work

Two other population-based studies have published home-time estimates [11,14]. A nation-wide Scottish study of over 101,000 patients defined home-time as we have done and reported estimates comparable to those observed here [14]. Home-time was not reported separately for haemorrhagic and ischaemic stroke and the gold-standard NIHSS severity measure was ascertained in around 5% of cases. Standardised outcome measures were unavailable. Another study identified ∼15,000 stroke cases representing all subtypes as here and counted overnight hospital stays only [11]. Validation of home-time was carried out for 552 patients with ischaemic stroke for whom the modified Rankin Scale scores were available.

In a multi-centre study of 1,866 subarachnoid haemorrhage patients with gold-standard case ascertainment, readmission data were not available [12]. Home-time strongly correlated with the modified Rankin Scale. Our results strengthen evidence for the validity of home-time in subarachnoid haemorrhage with more robust home-time assessment.

### 4.3 Strengths and limitations

Strengths include cost-effective, near universal home-time and proxy severity ascertainment, and population-based validation of home-time against baseline and outcome measures for all stroke sub-types with granular reporting of home-time by patient characteristics. Another strength was the capacity to capture same-day admissions including out-patient rehabilitation offered by hospitals and short-stay admissions as ignoring these admissions will artificially inflate home-time for patients with residual, ongoing disability.

Coding inaccuracy is a potential limitation although accuracy is often high [21]. As previously discussed [18], stroke coding in Australia has been validated in several studies showing high levels of accuracy in terms of positive predictive value, inter-rater coder reliability and sensitivity. Comorbidities are only coded if these require treatment in hospital and therefore it is likely comorbidities were under-numerated, although applying a look-back period reduces under-numeration [41]. The apparent protective effects of stroke history in subarachnoid haemorrhage and uncomplicated diabetes in intracerebral haemorrhage may reflect less severe aetiology (for example, non-aneurysmal aetiology for subarachnoid haemorrhage) or differential ascertainment of comorbidities in patients with dissimilar prognoses. Specifically, under-ascertainment may be more likely in patients who die within hours of admission or who are acutely unwell as the management of chronic comorbidities is not central to the care of critically ill patients. In contrast, comorbidities are relevant to relatively well patients and may be preferentially recorded in surviving patients.

FIM scores were obtained from in-patient rehabilitation records for the sub-group of patients receiving publicly funded rehabilitation who, by definition, had disability. These results may not generalise to patients undergoing rehabilitation as an outpatient or at private facilities. Home-time was not calculated for the 4.2% of patients either residing or transferred interstate or for those with missing hospitalisation data, and the impact of this source of missing data could not be assessed here. Anticipated national data linkage may eliminate shortcomings associated with cross-jurisdictional patient flows [42] while improved data quality will mitigate against data loss. We could not identify cases entering residential care who later returned to a private residence or patients admitted to supported care facilities after discharge to home, although the short 90-day follow-up period may have minimised such misclassification. We did not assess stroke severity using gold-standard measures as these are not available using administrative data, although the GCS is integral to measuring severity in subarachnoid haemorrhage [27] and is strongly predictive of mortality for ischaemic stoke [19, 26] and intracerebral haemorrhage [26].

Loss of consciousness may be an inexact proxy for severity. Very low GCS at first point of contact may indicate temporary loss of consciousness due to syncope or seizure, rather than stroke severity. This may account for the higher home-time estimates observed in haemorrhagic stroke patients – only in those surviving their admission – whose initial GCS scores fell within the most severe range compared with those whose scores were within the penultimate severity category. While GCS and mode of arrival predict mortality after stroke, the GCS is a weaker correlate of home-time than the National Institutes of Health Stroke Scale (NIHSS) [14], likely reflecting its less sensitive assessment of neurological deficits.

### 4.4 Implications for future research

Definitions of home-time have been adapted to suit the scope of administrative data sources at hand. For example, here and previously [14], home-time was defined as the number of full days not spent in hospital while others have calculated home-time as the number of nights spent at home [11]. Where available, residential care data provide reliable information about supportive care arrangements [11] whereas we and others [14] relied on hospital disposition coding to identify residential care admissions. Efforts are needed to harmonise the definition of home-time to aid multicentre and multinational studies and comparisons.

As noted by others [15], home-time may not accurately reflect disability for patients who receive assistance in their private residence nor does it differentiate between patients according to the level of support required in residential care. Patients discharged home who then die during the follow-up period may have equivalent home-time values as those who survive to 90-days with lengthy in-hospital stays. Further, discharge to nursing home care, death during the stroke admission and hospitalisation for the entire follow-up period yield equivalent home-time estimates of zero days. Whether the validity of home-time can be improved by adding qualitative information to achieve a more nuanced disability measure merits future research. Large differences in median values between the main cohort and the sub-set of patients surviving their stroke admission highlights the sizeable influence deaths have on home-time estimates. For this reason, we and others report descriptive analyses excluding in-patient deaths.

### 4.5 Conclusion

In conclusion, these findings support home-time ascertained using administrative data as a robust, inexpensive, replicable and valid patient-centred outcome that facilitates population-based stroke surveillance.

## Data Availability

Data analysed as part of this study are third-party owned. Signed agreements between the researchers and data custodians preclude sharing of individual level data in adherence to ethical and legal restrictions that apply to the use of these data-sets. Requests to access these third-party owned data-sets by qualified researchers can be directed to the Centre for Health Record Linkage (www.cherel.org.au). Requests for the programming code can be directed to the corresponding author.

## List of Abbreviations

IS: Ischaemic stroke
ICH: Intracerebral haemorrhage
SAH: Subarachnoid haemorrhage
NIHSS: National Institutes of Health Stroke Scale
FIM: Functional Independence Measure
GCS: Glasgow Coma Scale
NSW: New South Wales
eMR: electronic Medical Records (Ambulance NSW)
PHCR: Patient Health Care Record (Ambulance NSW)

## Acknowledgements

We thank the Centre for Health Record Linkage, for data linkage services and the NSW Ministry of Health and Ambulance NSW for supplying data for these analyses.

## Supplementary Material

**Supplementary Table 1:** The RECORD statement – checklist of items, extended from the STROBE statement, that should be reported in observational studies using routinely collected health data.

|  | Item No. | STROBE items | Location in manuscript where items are reported | RECORD items | Location in manuscript where items are reported |
| --- | --- | --- | --- | --- | --- |
| **Title and abstract** |  |  |  |  |  |
|  | 1 | (a) Indicate the study's design with a commonly used term in the title or the abstract (b) Provide in the abstract an informative and balanced summary of what was done and what was found | Title, Abstract | RECORD 1.1: The type of data used should be specified in the title or abstract. When possible, the name of the databases used should be included.<br>RECORD 1.2: If applicable, the geographic region and timeframe within which the study took place should be reported in the title or abstract.<br>RECORD 1.3: If linkage between databases was conducted for the study, this should be clearly stated in the title or abstract. | Title (administrative data is specified).<br>Abstract, Line 43.<br>Title; Abstract, lines 43-44. |
| **Introduction** |  |  |  |  |  |
| Background rationale | 2 | Explain the scientific background and rationale for the investigation being reported | Introduction, Lines 64-98. |  | Introduction, Lines 64-98. |
| Objectives | 3 | State specific objectives, including any prespecified hypotheses | Introduction, Lines 92-98. |  |  |

| Methods |  |  |  |  |  |
| --- | --- | --- | --- | --- | --- |
| Study Design | 4 | Present key elements of study design early in the paper | Methods, Lines 102-111 |  |  |
| Setting | 5 | Describe the setting, locations, and relevant dates, including periods of recruitment, exposure, follow-up, and data collection | Methods, Lines 112-120. |  |  |
| Participants | 6 | (a) *Cohort study* - Give the eligibility criteria, and the sources and methods of selection of participants. Describe methods of follow-up<br>*Case-control study* - Give the eligibility criteria, and the sources and methods of case ascertainment and control selection. Give the rationale for the choice of cases and controls<br>*Cross-sectional study* - Give the eligibility criteria, and the sources and methods of selection of participants<br>(b) *Cohort study* - For matched studies, give matching criteria and number of exposed and unexposed<br>*Case-control study* - For matched studies, give matching criteria and the number of controls per case | Not applicable: this is neither a cohort nor a case-control study<br>Not applicable: this is neither a cohort nor a case-control study. | RECORD 6.1: The methods of study population selection (such as codes or algorithms used to identify subjects) should be listed in detail. If this is not possible, an explanation should be provided.<br>RECORD 6.2: Any validation studies of the codes or algorithms used to select the population should be referenced. If validation was conducted for this study and not published elsewhere, detailed methods and results should be provided.<br>RECORD 6.3: If the study involved linkage of databases, consider use of a flow diagram or other graphical display to demonstrate the data linkage process, including the number of individuals with linked data at each stage. | Methods, lines 112-141.<br>Methods, Lines 115-127; citation to relevant references 18,19,21,22<br>Figure 1. |
| Variables | 7 | Clearly define all outcomes, exposures, predictors, potential confounders, and effect modifiers. Give diagnostic criteria, if applicable. | Methods, Lines 142-151 (outcome described); Lines 162-175, incorporating reference 19 describing previous validation; Lines 176 to 185; Lines 203 to 207. | RECORD 7.1: A complete list of codes and algorithms used to classify exposures, outcomes, confounders, and effect modifiers should be provided. If these cannot be reported, an explanation should be provided. | Methods, Lines 142-151 (outcome described); Lines 162-175, incorporating reference 19 describing previous validation; Lines 176 to 185; Lines 203 to 207. |
| Data sources/<br>measurement | 8 | For each variable of interest, give sources of data and details of methods of assessment (measurement). Describe comparability of assessment methods if there is more than one group | Methods, Lines 142-151 (outcome described); Lines 162-175, incorporating reference 19 describing previous validation; Lines 176 to 185; Lines 203 to 207. |  |  |
| Bias | 9 | Describe any efforts to address potential sources of bias | Lines 132 to 141. |  |  |
| Study size | 10 | Explain how the study size was arrived at | Methods, line 112 to 114 (all available cases) |  |  |
| Quantitative variables | 11 | Explain how quantitative variables were handled in the analyses. If applicable, describe which groupings were chosen, and why | Methods, Statistical analysis. Lines 186 to 192; Lines 210 to 216. |  |  |
| Statistical methods | 12 | (a) Describe all statistical methods, including those used to control for confounding<br>(b) Describe any methods used to examine subgroups and interactions<br>(c) Explain how missing data were addressed<br>(d) *Cohort study* - If applicable, explain how loss to follow-up was addressed<br>*Case-control study* - If applicable, explain how matching of cases and controls was addressed<br>*Cross-sectional study* - If applicable, describe analytical methods taking account of sampling strategy<br>(e) Describe any sensitivity analyses | Methods, Statistical Analysis, Lines 186 to 217.<br>Figure 1.<br>Not applicable.<br>Not applicable<br>Not applicable<br>e) Lines 189-192. |  | . |
| Data access and cleaning methods |  | .. |  | RECORD 12.1: Authors should describe the extent to which the investigators had access to the database population used to create the study population.<br>RECORD 12.2: Authors should provide information on the data cleaning methods used in the study. | Methods, Line 104 to 111.<br>Data statement, Lines 499 to 503.<br>Methods, Line |
| Linkage |  | .. |  | RECORD 12.3: State whether the study included person-level, institutional-level, or other data linkage across two or more databases. The methods of linkage and methods of linkage quality evaluation should be provided. | Figure 1, Lines 105 to 111. |
| **Results** |  |  |  |  |  |
| Participants | 13 | (a) Report the numbers of individuals at each stage of the study (*e.g.*, numbers potentially eligible, examined for eligibility, confirmed eligible, included in the study, completing follow-up, and analysed)<br>(b) Give reasons for non-participation at each stage.<br>(c) Consider use of a flow diagram | a) Figure 1.<br>b) Lines 222 to 228.<br>c) Figure 1. | RECORD 13.1: Describe in detail the selection of the persons included in the study (*i.e.*, study population selection) including filtering based on data quality, data availability and linkage. The selection of included persons can be described in the text and/or by means of the study flow diagram. | Figure 1, Lines 112 to 136. |
| Descriptive data | 14 | (a) Give characteristics of study participants (*e.g.*, demographic, clinical, social) and information on exposures and potential confounders<br>(b) Indicate the number of participants with missing data for each variable of interest<br>(c) *Cohort study* - summarise follow-up time (*e.g.*, average and total amount) | a) Supplementary Tables 2-5.<br>b) Figure 1, Supplementary Tables 2-5.<br>c) Not applicable |  |  |
| Outcome data | 15 | *Cohort study* - Report numbers of outcome events or summary measures over time | Tables 1 and 2. |  |  |
|  |  | *Case-control study* - Report numbers in each exposure category, or summary measures of exposure<br>*Cross-sectional study* - Report numbers of outcome events or summary measures |  |  |  |
| Main results | 16 | (a) Give unadjusted estimates and, if applicable, confounder-adjusted estimates and their precision (e.g., 95% confidence interval). Make clear which confounders were adjusted for and why they were included<br>(b) Report category boundaries when continuous variables were categorized<br>(c) If relevant, consider translating estimates of relative risk into absolute risk for a meaningful time period | Tables 1 to 3. Supplementary Tables 2 to 5. |  |  |
| Other analyses | 17 | Report other analyses done—e.g., analyses of subgroups and interactions, and sensitivity analyses | Tables 1 to 3 (removing patients who died in hospital). |  |  |
| **Discussion** |  |  |  |  |  |
| Key results | 18 | Summarise key results with reference to study objectives | Lines 361 to 405. |  |  |
| Limitations | 19 | Discuss limitations of the study, taking into account sources of potential bias or imprecision. | Lines 419 to 460 | RECORD 19.1: Discuss the implications of using data that were not created or collected to answer the specific research question(s). Include | Lines 426 to 460. Lines 461 to 480. |
|  |  | Discuss both direction and magnitude of any potential bias |  | discussion of misclassification bias, unmeasured confounding, missing data, and changing eligibility over time, as they pertain to the study being reported. |  |
| Interpretation | 20 | Give a cautious overall interpretation of results considering objectives, limitations, multiplicity of analyses, results from similar studies, and other relevant evidence | Discussion. |  |  |
| Generalisability | 21 | Discuss the generalisability (external validity) of the study results | Lines 406 to 418; 461 to 480. |  |  |
| **Other Information** |  |  |  |  |  |
| Funding | 22 | Give the source of funding and the role of the funders for the present study and, if applicable, for the original study on which the present article is based | Lines 487 to 492. |  |  |
| Accessibility of protocol, raw data, and programming code |  | .. |  | RECORD 22.1: Authors should provide information on how to access any supplemental information such as the study protocol, raw data, or programming code. | Lines 499 to 504. |

\*Reference: Benchimol EI, Smeeth L, Guttman A, Harron K, Moher D, Petersen I, Sørensen HT, von Elm E, Langan SM, the RECORD Working Committee. The REporting of studies Conducted using Observational Routinely-collected health Data (RECORD) Statement. *PLoS* *Medicine* 2015; in press.
\*Checklist is protected under Creative Commons Attribution (CC BY) license.

**Supplementary Table 2:** Patient characteristics (N=74,501)

| Characteristics | N | % |
| --- | --- | --- |
| Sub-type |  |  |
| Ischaemic Stroke | 57,949 | 77.8 |
| Intracerebral Haemorrhage | 12,192 | 16.4 |
| Subarachnoid Haemorrhage | 4,360 | 5.9 |
| Sex |  |  |
| Male | 37,404 | 50.2 |
| Female | 37,097 | 49.8 |
| Age at diagnosis (years) |  |  |
| <50 | 5,205 | 7.0 |
| 50-64 | 11,520 | 15.5 |
| 65-74 | 14,692 | 19.7 |
| 75-84 | 24,731 | 33.2 |
| 85+ | 18,353 | 24.6 |
| Stroke Severity (IS and ICH);<br>N=70,141** |  |  |
| Ambulant | 23,751 | 33.9 |
| GCS score=15 | 25,429 | 36.3 |
| GCS score=13-14 | 9,001 | 12.8 |
| GCS score=9-12 | 7,219 | 10.3 |
| GCS score=3-8 | 4,215 | 6.0 |
| Missing* | 526 | 0.7 |
| Stroke Severity (SAH);<br>N=4,360** |  |  |
| Ambulant | 1,600 | 36.7 |
| GCS score=15 | 1,297 | 29.7 |
| GCS score=13-14 | 392 | 9.0 |
| GCS score=7-12 | 459 | 10.5 |
| GCS score=3-6 | 585 | 13.4 |
| Missing* | 27 | 0.6 |

\*Patients arriving by ambulance with missing GCS (Glasgow Coma Scale) scores
\*\*Denominator used for calculation of percentages

**Supplementary Table 3:**
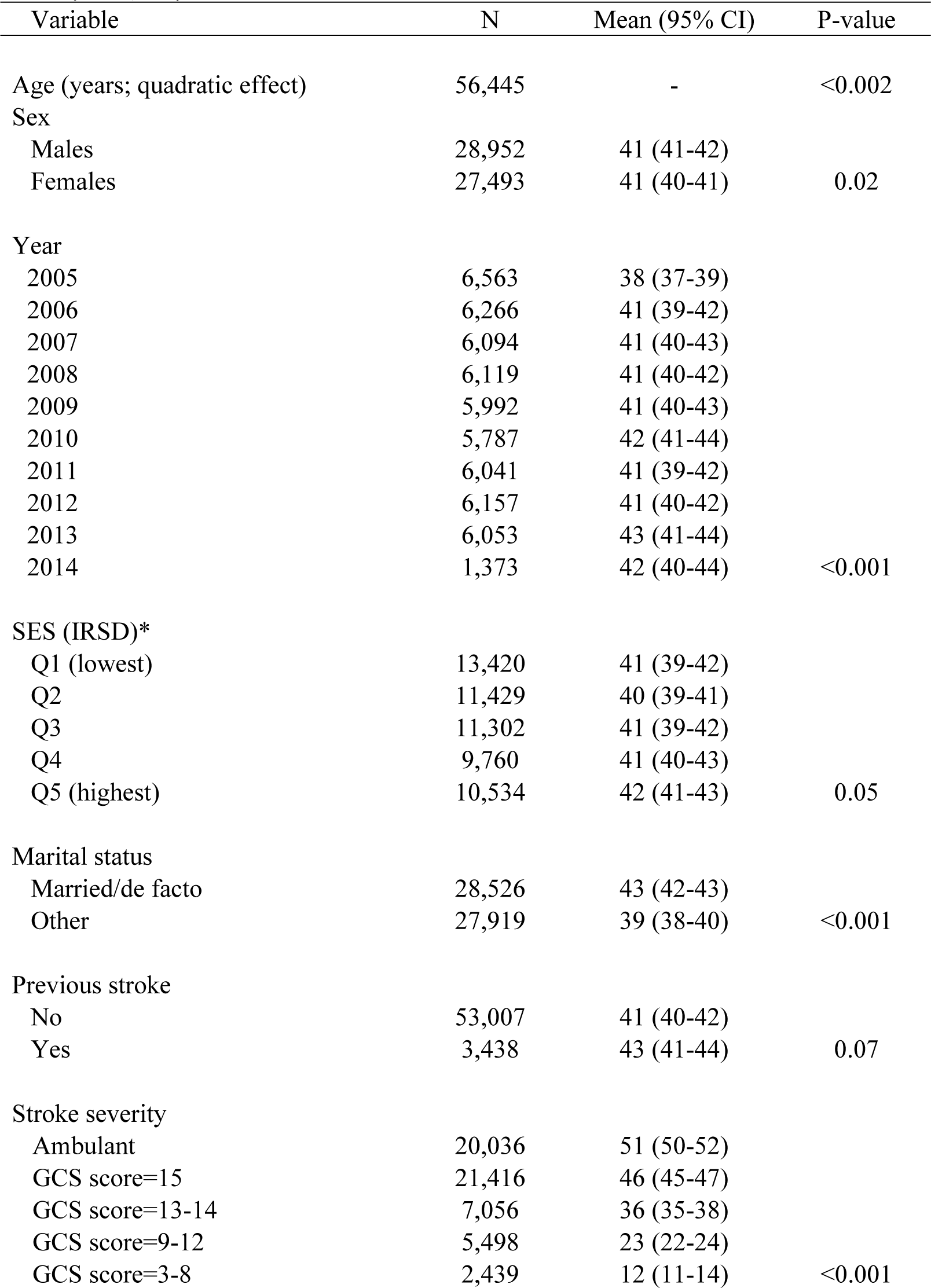
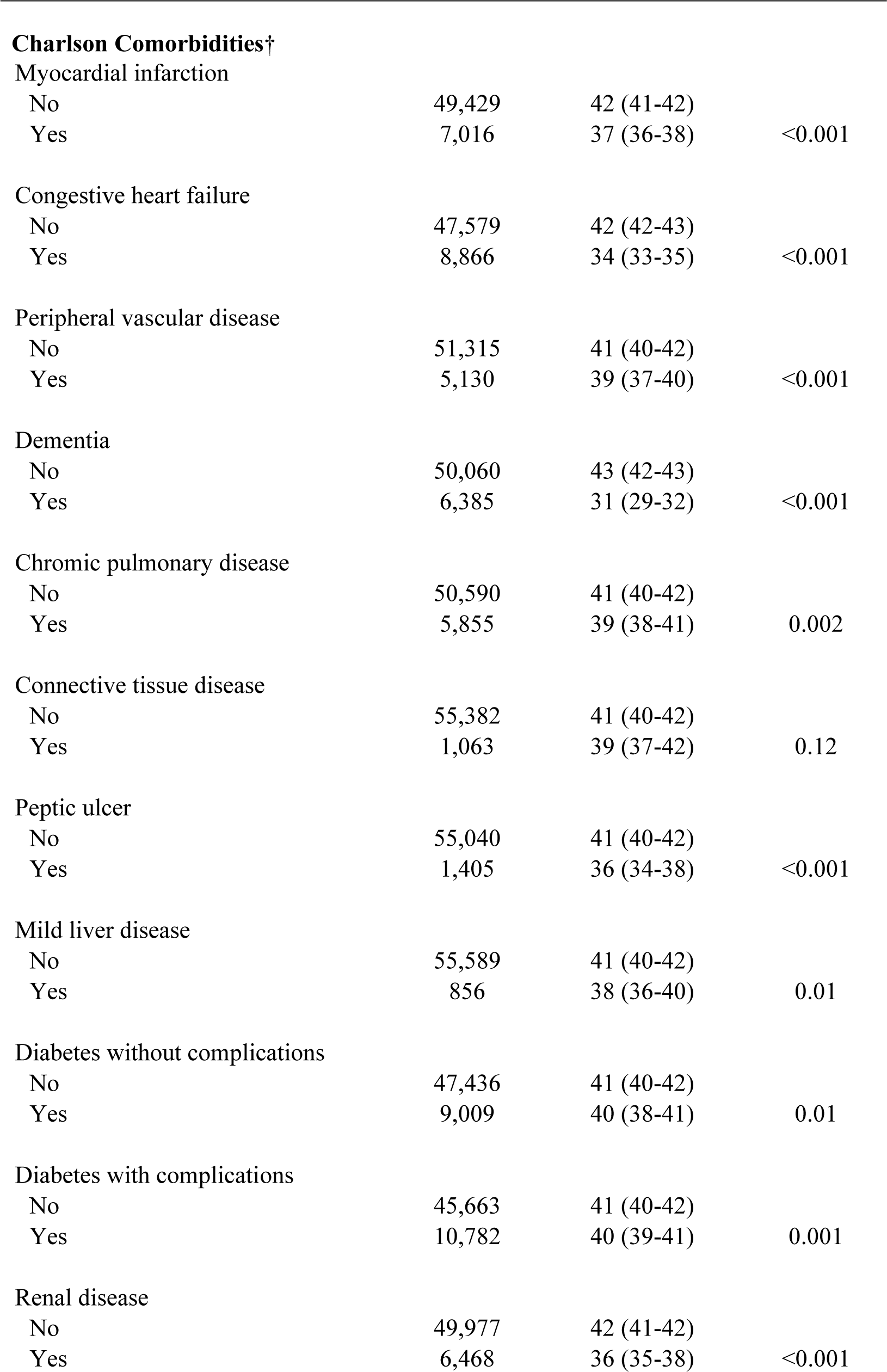
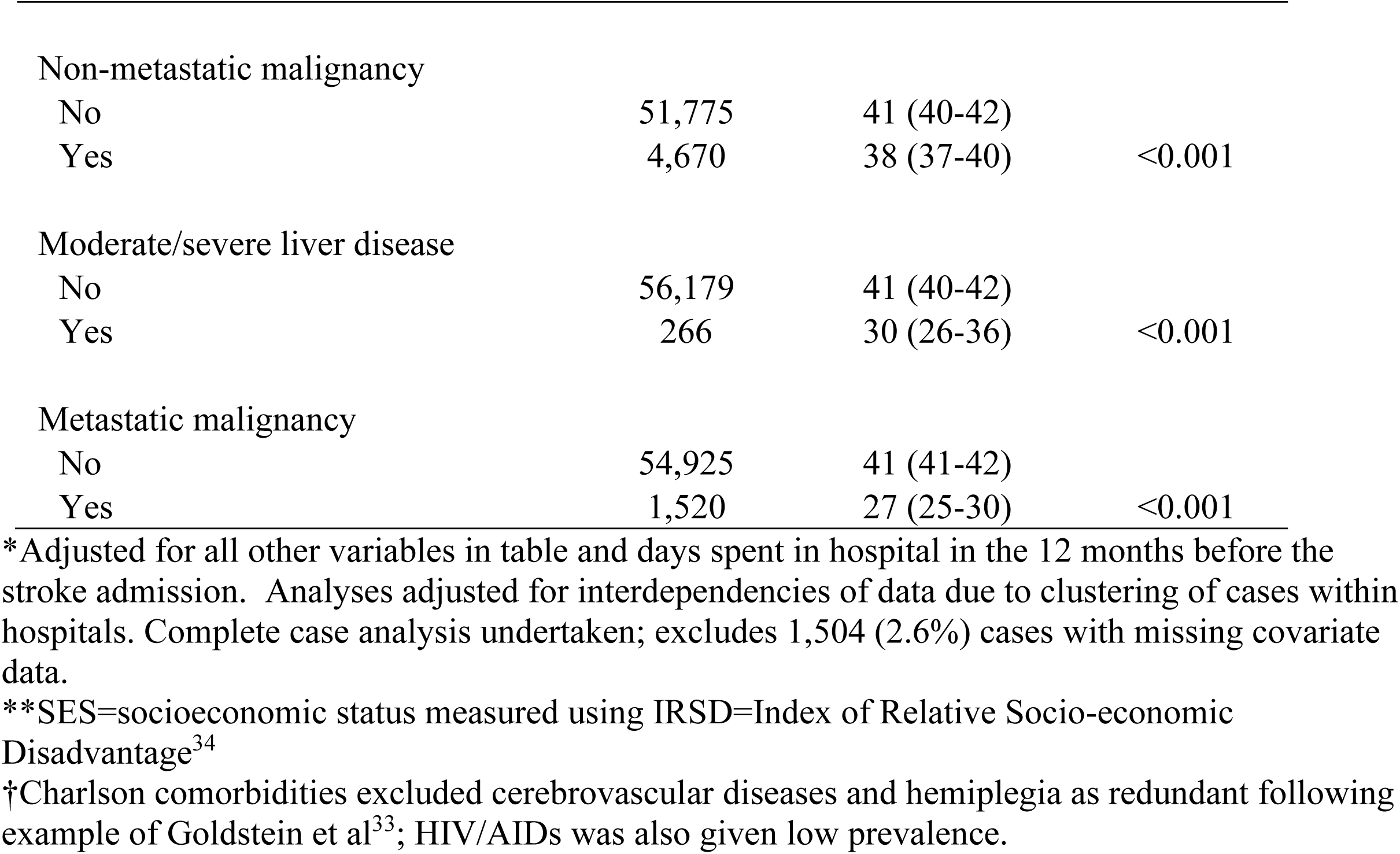
Adjusted* mean days spent at home: Ischaemic Stroke (n=56,445)*

**Supplementary Table 4:**
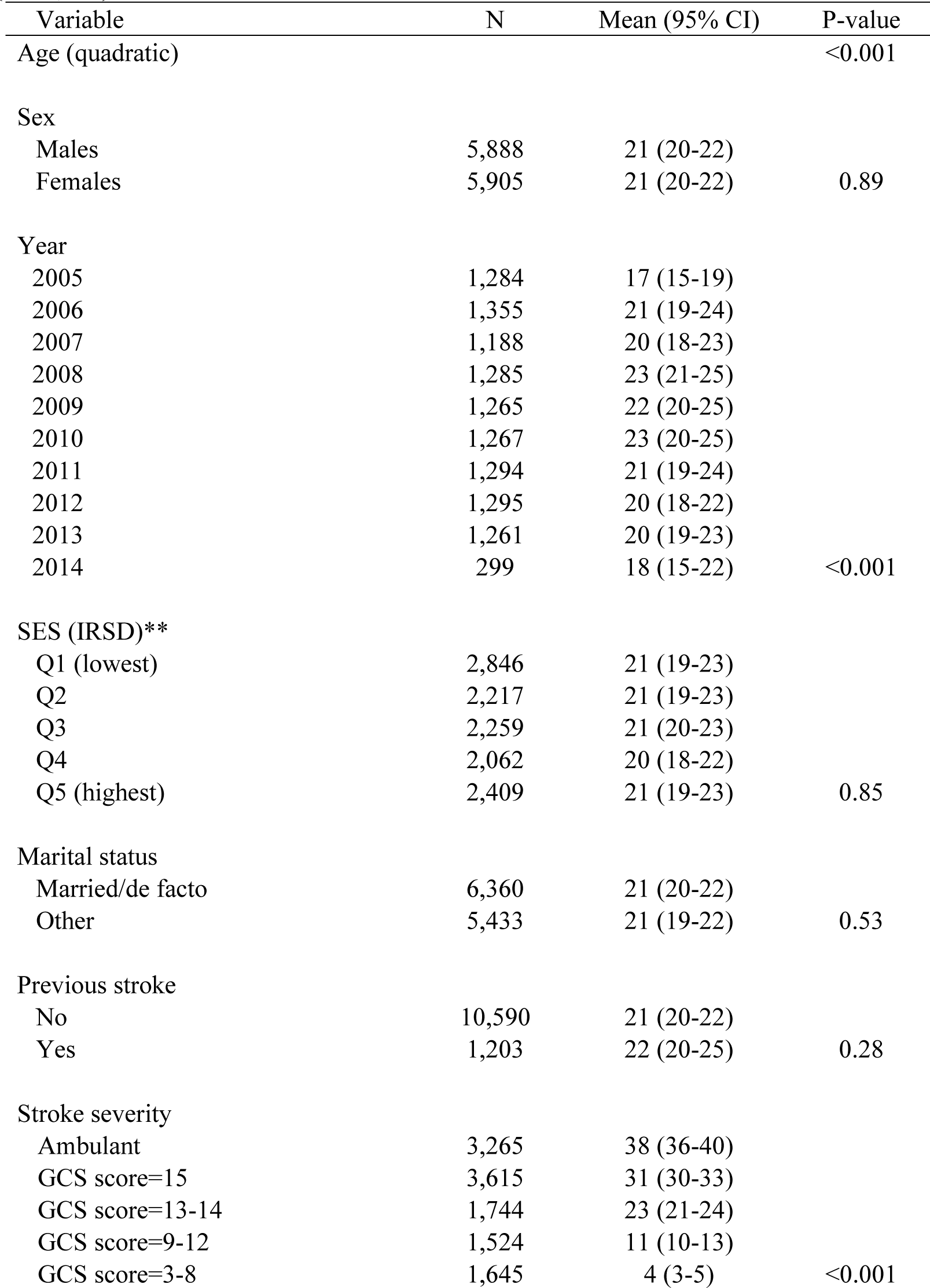
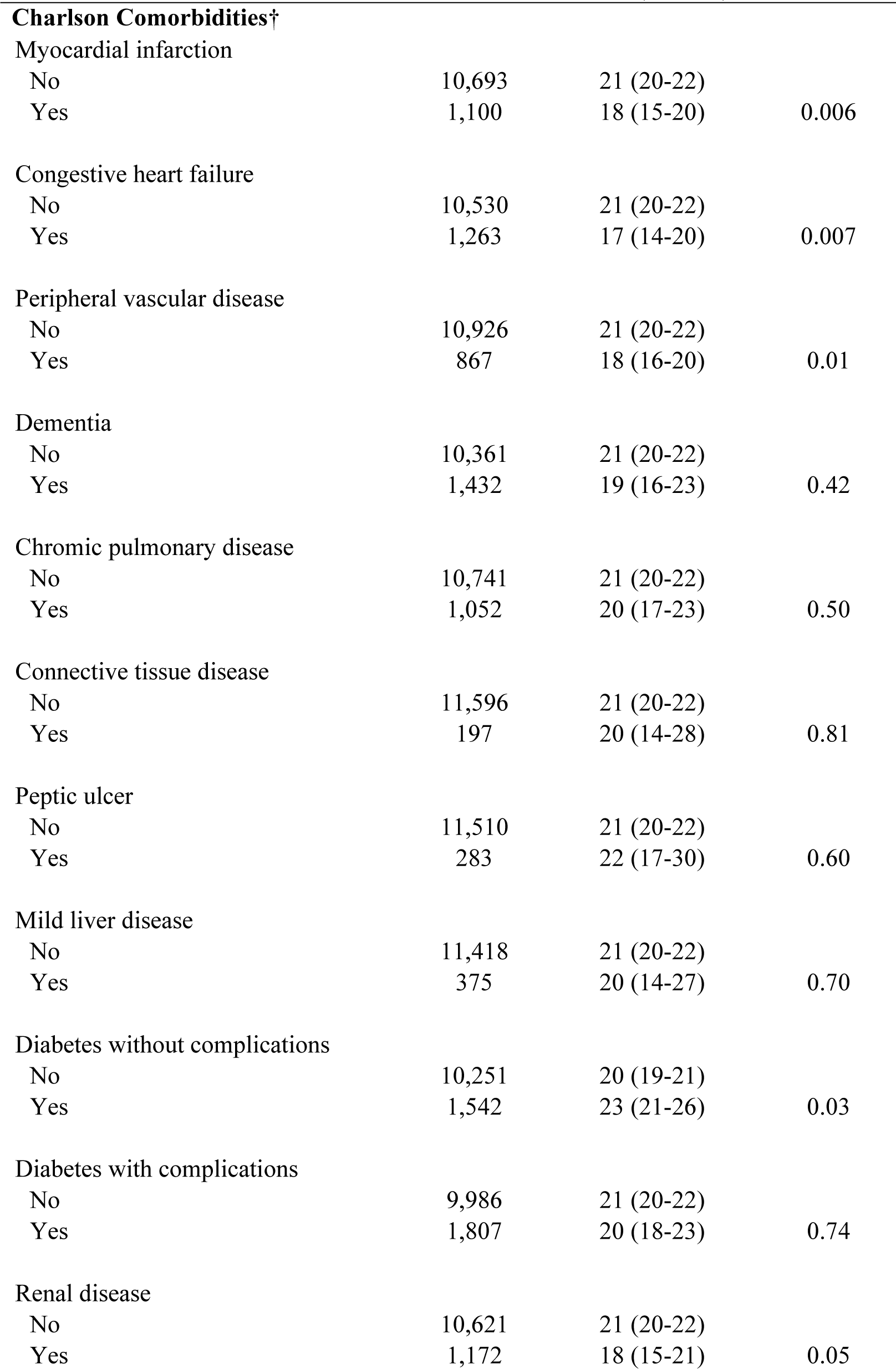
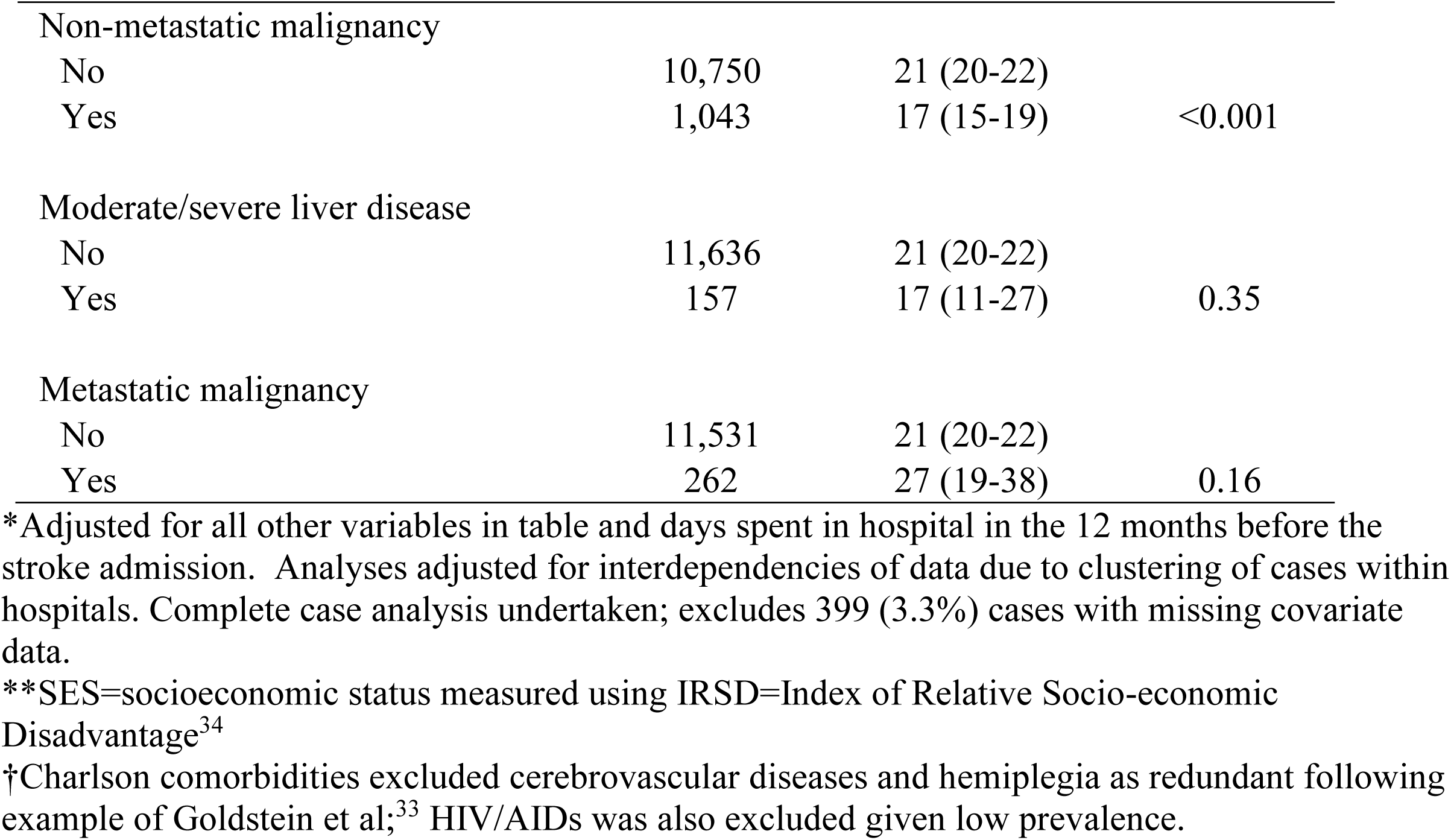
Adjusted* mean days spent at home: ICH (n=11,793)

**Supplementary Table 5:**
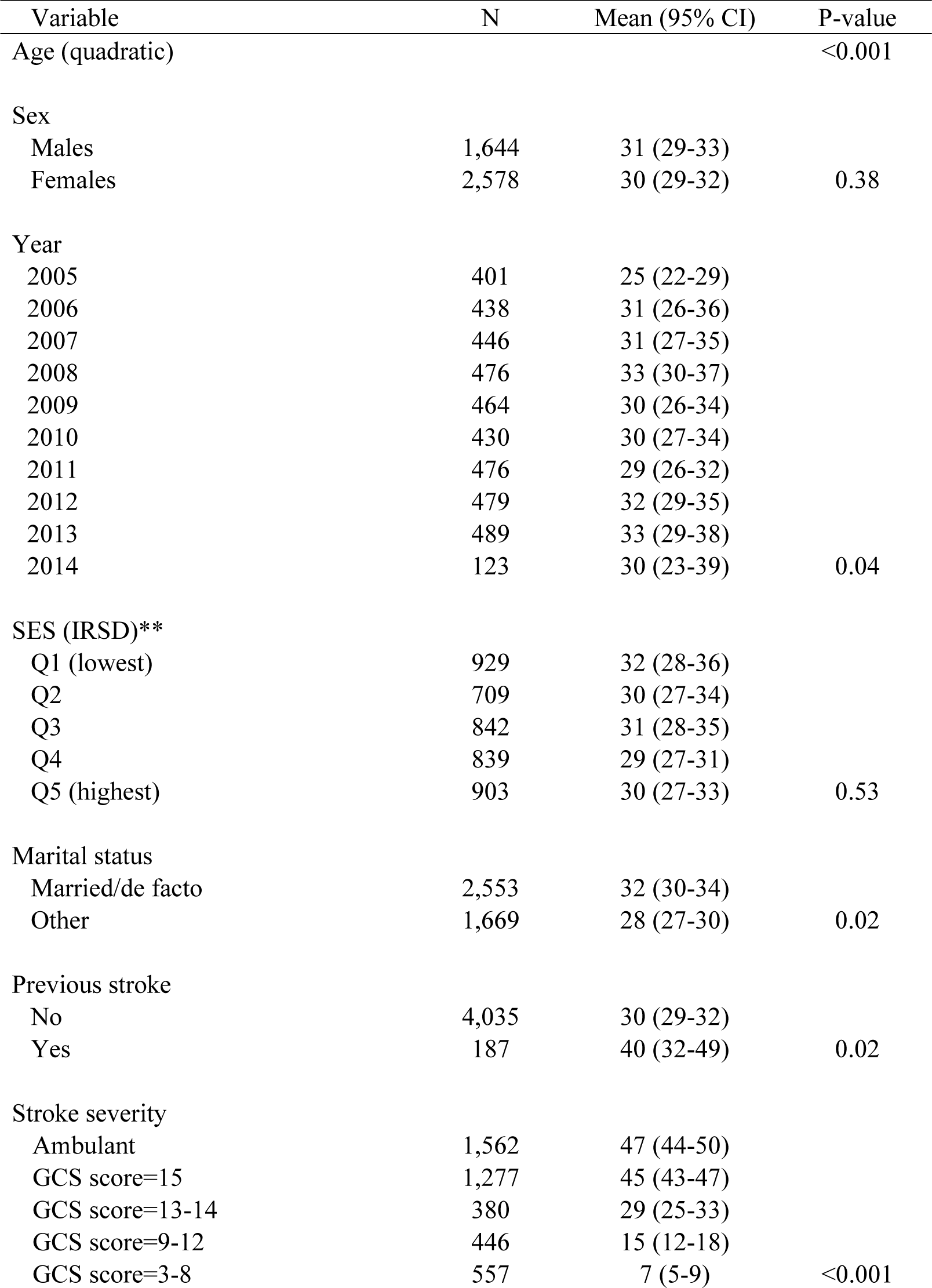
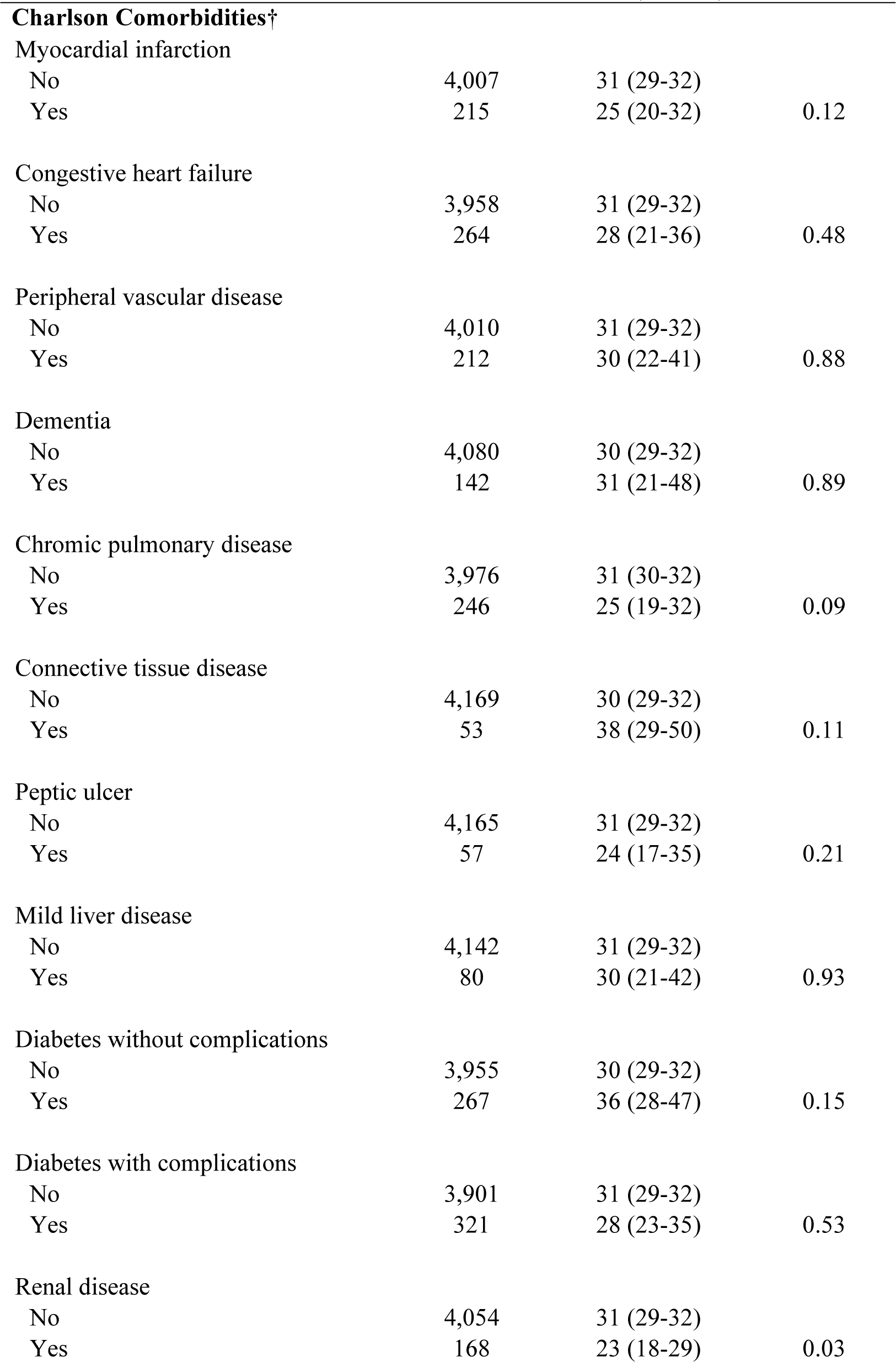
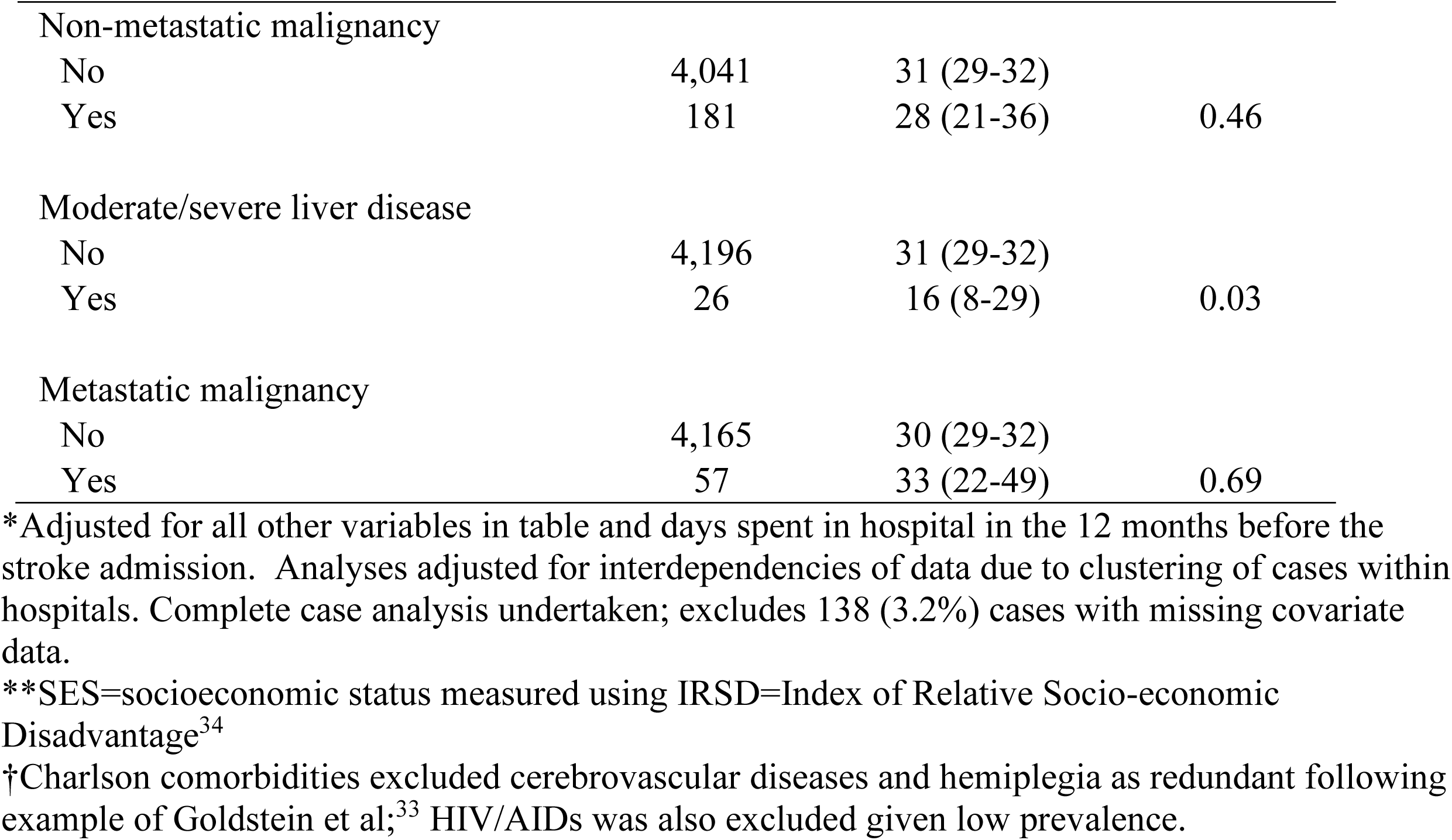
Adjusted* mean days spent at home: SAH (n=4,222)

